# The Effect of Low Back Pain Daoyin Exercise Based on the Theory of Goal Attainment on Patients with Lumbar Disc Herniation: Study protocol of a randomized controlled trial

**DOI:** 10.64898/2026.03.17.26348594

**Authors:** Mutong Wang, Shuai Guo, Yican Yang, Gengyu Liang, Zishan Wang, Yang Zhang

## Abstract

**Background:** The prevalence of lumbar disc herniation (LDH) is increasing, and the associated pain and functional limitations severely impact patients’ quality of life. Daoyin, a traditional Chinese exercise, has a history of thousands of years in managing musculoskeletal pain. However, its application in LDH has not been sufficiently investigated, and there is a notable scarcity of rigorous randomized controlled trials (RCTs). This paper outlines the protocol for an RCT based on the theory of goal attainment (TGA), which aims to investigate whether Daoyin is more effective than other exercise therapies in improving symptoms in patients with LDH.

**Methods:** We conducted a 6-week RCT in which participants were randomly assigned to either Daoyin or core stability exercise (CSE). During the first two weeks, the participants performed their assigned exercises five times per week. Outcome data were collected at baseline, week 2, and week 6. The primary outcome was pain intensity at 6 weeks, which was assessed via the visual analogue scale (VAS). The secondary outcomes included the Japanese Orthopaedic Association (JOA) score, the Oswestry Disability Index (ODI), the MOS 36-item Short Form Health Survey (SF-36), the Hospital Anxiety and Depression Scale (HADS), surface electromyography (sEMG), gait analysis, and electroencephalography (EEG). A generalized estimating equation (GEE) model will be used to analyse longitudinal changes and between-group differences.

**Discussion:** This trial seeks to assess the efficacy of Daoyin for LDH and to elucidate its underlying neuromuscular mechanisms. Should the intervention prove feasible, the findings will inform the design of a subsequent large-scale RCT and are expected to contribute to a solid evidence base for the broader clinical application of Daoyin.

**Trial registration:** https://itmctr.ccebtcm.org.cn/, Registration number (ITMCTR2025001239).

## Introduction

Lumbar disc herniation (LDH) commonly manifests as low back pain (LBP), radiating neuropathic pain in the lower limbs, numbness, and restricted lumbar mobility[1]. In recent years, changes in work and lifestyle have led to a sharp increase in the number of LDH patients, with a trend toward younger onset. This condition significantly impairs patients’ physical and mental health, as well as their quality of life, making it a major threat to global health[2].

LBP is the most frequent symptom among LDH patients and a leading cause of productivity loss and disability worldwide[1,3]. Owing to its persistent and often progressive nature, LBP imposes substantial psychological and economic burdens on individuals[3,4]. The World Health Organization has identified the reduction of disabling LBP as a global health goal and advocates that LBP-related research be prioritized in global health initiatives[4].

Nonsurgical treatments are widely used in clinical practice to alleviate LBP. However, commonly prescribed medications—such as nonsteroidal anti-inflammatory drugs, muscle relaxants, and tricyclic antidepressants—often provide only temporary pain relief and are associated with potential side effects[1]. Clinical practice guidelines recommend exercise as a first-line therapy for LBP[5,6]. Exercise therapies, including core stability exercise (CSE), Pilates, yoga, and Tai Chi, have been shown to effectively reduce pain severity, enhance daily functioning, and improve mood in LBP patients[7–9].

Daoyin, a traditional Chinese movement therapy, has demonstrated potential in managing LBP[10]. Practices such as Baduanjin, Wuqinxi, and Tai Chi fall under the category of Daoyin. This approach integrates breathing techniques with physical movements to regulate respiration and mental focus, promoting qi and blood circulation, muscle relaxation, and smooth movement[10]. Although several studies have applied Daoyin in LBP treatment, the existing evidence is limited by low methodological quality and a lack of objective outcome measures, which undermines its utility for clinical decision-making [10].

The Theory of Goal Attainment (TGA) is an interactive framework designed to facilitate behavioral change in patients[11]. It consists of five stages: interactive assessment, diagnosis, planning, implementation, and evaluation[12]. The assessment comprehensively addresses personal systems (e.g., self, body image, perception, growth), interpersonal systems (e.g., communication, interaction, role), and social systems (e.g., organization, authority, status)[12]. The TGA emphasizes nurse–patient communication, uses interaction to identify issues, enhances patient awareness, and collaboratively sets health-related goals and plans [12]. The conceptual framework of the TGA is shown in Fig 1. By focusing on communication, goal setting, and patient-centered care, TGA can enhance patient adherence and facilitate the implementation of effective interventions [11]. To date, TGA has been successfully applied in conditions such as multiple sclerosis, type 2 diabetes, and acute coronary syndrome, improving self-management, adherence, functional capacity, and quality of life[13,14]. However, its application in LBP remains limited.

**Fig 1.**
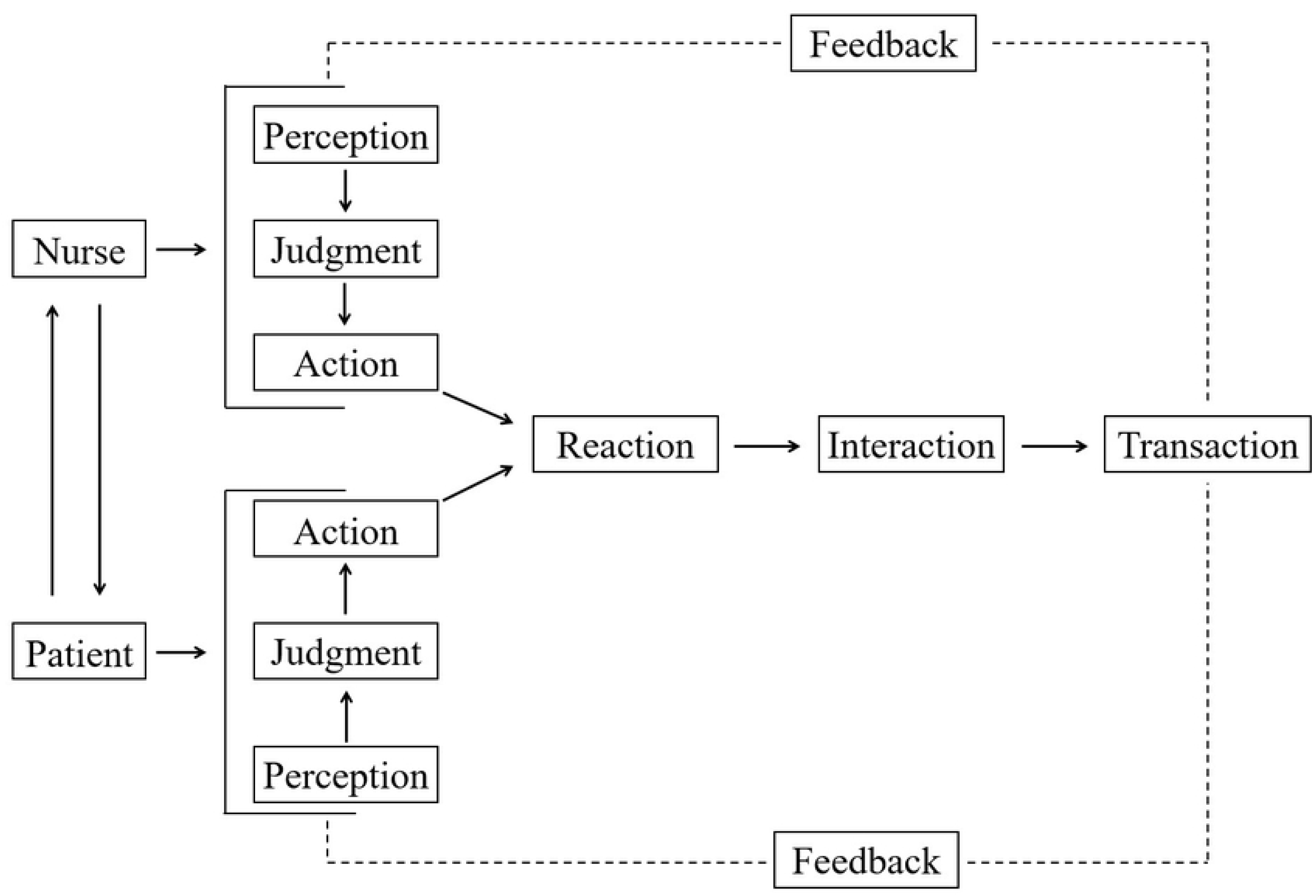
TGA Conceptual Framework. This figure illustrates the interactive process between the nurse and the patient. Both parties move through perception, judgment, and action, which lead to reaction, interaction, and transaction, with feedback operating throughout the whole process. Solid arrows indicate the direction of progression, the bidirectional arrow indicates mutual influence between nurse and patient, and dashed lines indicate feedback loops.

Therefore, this study adopts TGA to assess patients with LDH and establish individualized Daoyin exercise goals and plans. A randomized controlled trial (RCT) will be conducted to evaluate the efficacy and safety of Daoyin for LBP in patients with LDH.

## Methods

### Study design

This study employs a randomized, controlled, single-blind clinical trial to evaluate the efficacy of Daoyin for LBP in patients with LDH. Participants will be recruited from Shandong University of Traditional Chinese Medicine Affiliated Hospital and the surrounding communities using recruitment posters. Eligible participants will be randomly assigned in a 1:1 ratio to the Daoyin group or the CSE group. This study began recruiting participants on 6 July 2025 and is currently ongoing, with recruitment planned to conclude in May 2026. All participants provided written informed consent. The total study duration was 6 weeks, which included 2 weeks of intervention training. Assessments will be conducted at baseline (T0), week 2 (T1), and week 6 (T2).

The primary outcome was the change in pain intensity at 6 weeks, which was measured via the visual analogue scale (VAS). The secondary outcomes included evaluations of pain, lumbar function, quality of life, and emotional status. These will be quantified via the following instruments: electroencephalography (EEG), the Japanese Orthopaedic Association (JOA), the Oswestry Disability Index (ODI), surface electromyography (sEMG), three-dimensional gait analysis, the MOS 36-item Short Form Health Survey (SF-36), and the Hospital Anxiety and Depression Scale (HADS). These measures will help determine whether Daoyin is noninferior or superior to CSE in efficacy.

The trial will be conducted at the Department of Rehabilitation and Physiotherapy, The Affiliated Hospital of Shandong University of Traditional Chinese Medicine. The study protocol was approved by the Ethics Committee of the School of Nursing and Rehabilitation, Shandong University (Approval No: 2024-R-155; Approval date: December 31, 2024). It has been registered with the International Traditional Medicine Clinical Trial Registry (Registration No: ITMCTR2025001239; Registered on 21 June 2025).

The study protocol adheres to the Standard Protocol Items: Recommendations for Interventional Trials (SPIRIT) guidelines (The detailed checklist is provided in S1 Table). The study design is summarized in the flow diagram presented in Fig 2. A summary of the study schedule, which includes participant enrollment, interventions, and assessments, is presented in Table 1.

**Fig 2.**
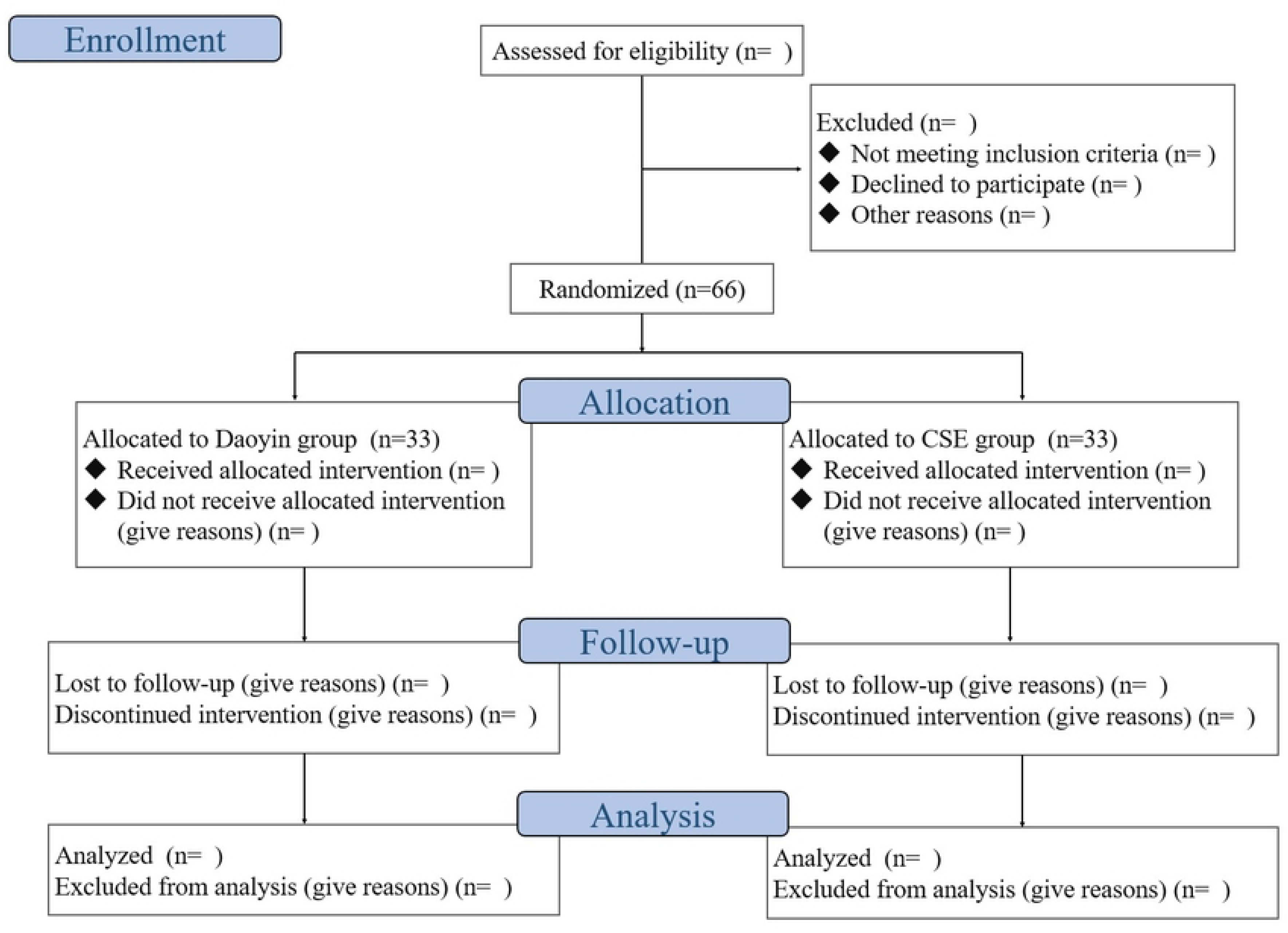
Study Design Flowchart. This figure depicts the study process, from eligibility assessment and randomization through allocation, follow-up, and analysis. A total of 66 participants were randomized to the Daoyin group (n=33) or the Core Stability Exercise (CSE) group (n=33). The diagram also reports the numbers of exclusions, loss to follow-up, discontinuation, and final analysis at each stage.

**Table 1.** SPIRIT 2025 participant timeline.

|  | TRIAL PERIOD |  |  |  |
| --- | --- | --- | --- | --- |
|  | Enrollment |  | Post-randomization | Close-out |
| TIMEPOINT | $-t_1$ to 0 | 0 | $t_1$ | $t_2$ |
| ENROLLMENT: |  |  |  |  |
| Eligibility screen | X |  |  |  |
| Exclusion criteria | X |  |  |  |
| Informed consent | X |  |  |  |
| Randomization |  | X |  |  |
| INTERVENTION/<br>COMPARATOR: |  |  |  |  |
| TGA-based LBP Daoyin<br>exercisec |  | X | → | X |
| CSE |  | X | → | X |
| ASSESSMENTS: |  |  |  |  |
| Demographic<br>characteristics |  | X |  |  |
| VAS |  | X | X | X |
| ODI |  | X | X | X |
| JOA |  | X | X | X |
| gait analysis |  | X | X | X |
| sEMG |  | X | X | X |
| EEG |  | X | X | X |
| SF-36 |  | X | X | X |
| HADS |  | X | X | X |

### Sample size

The sample size calculation was based on the hypothesized change in the VAS score. For the primary outcome of change in pain intensity from baseline to week 6, a two-sided alpha error of 0.05 and a power of 0.9 were assumed. On the basis of previous literature, the effect size for the VAS score was estimated to be 0.86[15]. The total sample size was calculated via G*Power software (version 3.1, Germany). Accounting for a 10% dropout rate, a minimum of 66 participants are needed to detect a statistically significant clinical effect of the 6-week Daoyin intervention compared with CSE in patients with LDH.

### Inclusion criteria

Eligible participants must meet all of the following criteria: (i) were diagnosed with LDH confirmed by CT or MRI and physical examination; (ii) were in the chronic phase of LDH; (iii) presented with LBP symptoms; (iv) were aged between 18 and 75 years; (v) were proficient in using a smartphone; (vi) did not have significant visual or auditory impairments that would hinder the ability to learn Daoyin movements; and (vii) provided informed consent and voluntarily participated in the study.

### Exclusion criteria

Participants were excluded based on any of the following criteria: (i) LBP attributed to other causes, such as lumbar muscle strain, sprain, or urological disorders; (ii) comorbid spinal conditions, including lumbar spinal stenosis, spondylolisthesis, spinal tuberculosis, or spinal tumors; (iii) participation in other traditional Chinese exercise therapies (e.g., Yijinjing, Wuqinxi, Liuzijue) within the past three months; or (iv) severe primary diseases or unstable conditions, including significant dysfunction of the heart, liver, spleen, or kidneys; autoimmune diseases; or malignant tumors.

### Withdrawal and Dropout Criteria

Participants may withdraw from the study at any time for any reason, provided that they do so voluntarily. Additionally, participants will be discontinued from the study in cases of poor adherence (e.g., failure to respond to two consecutive follow-up calls), disease progression, or the onset of other severe medical conditions.

### Randomization and Allocation

Eligible participants will be randomly assigned in a 1:1 ratio to either the Daoyin group or the CSE group. Randomization will be performed by an independent researcher using a computer-generated sequence created with IBM SPSS software (Version 27.0).To maintain the confidentiality of the allocation, allocation information will be securely sealed in opaque envelopes bearing sequential serial numbers. Upon completion of subject registration, an independent researcher will sequentially draw the envelopes and communicate the allocation information to the intervention implementers.

### Blinding

A single-blind trial design was implemented. To minimize bias, blinding was applied to the study recruiters, participants, outcome assessors, and data analysts. However, the personnel administering the interventions were necessarily unblinded due to the nature of the intervention. To protect the blinding of participants, the exercise regimens for both groups were matched not only for the number of movements and total session duration but also for similar starting postures.

### Interventions

#### Daoyin group

Implementation of the TGA-based LBP Daoyin exercise intervention protocol, which has undergone two rounds of expert consultation via the Delphi method. The detailed protocol is provided in Supplementary Material 2. It consists of five structured movements, including “Rowing with Both Elbows” and “Xianfengzhangchi.” Under the guidance of an experienced Daoyin instructor, participants will engage in 30-minute sessions five times per week for two weeks. Following this training phase, participants will be provided with instructional videos to support independent home practice, which they are advised to perform at least three times per week. Participants can provide exercise feedback to coaches via WeChat to monitor adherence.

#### CSE group

Multiple studies have demonstrated that CSE can significantly alleviate pain and functional disability in patients with LDH or chronic LBP, while enhancing muscle strength and improving dynamic spinal stability[16,17]. Thus, employing CSE as a control intervention is consistent with evidence-based medicine and ensures that the control group receives an effective treatment. The participants in this group will perform CSE under the guidance of a senior physiotherapist for 30 minutes per session, five times weekly over two weeks. The CSE protocol comprises five exercises: Cat-Cow Stretch, Modified Plank, Glute Bridge, Dead Bug, and Bird-Dog. The specific procedures for each exercise are detailed in Supplementary Material 2. Instructional videos were also provided after the training period, and one structured telephone follow-up was conducted after the two-week training phase.

#### Strategies to Enhance Adherence to Interventions

Before trial initiation, participants were informed about the potential benefits of Daoyin exercise or CSE in alleviating low back pain to enhance their understanding of the trial’s value and to foster active engagement. During the intervention period, individualized one-to-one instruction was provided to ensure accurate performance of the prescribed movements, with timely feedback and encouragement to strengthen participants’ sense of achievement. In addition, follow-up was conducted via telephone or WeChat to proactively identify any difficulties encountered by participants and adjust the intervention plan as needed.

### Data collection

All data will be assessed at three time points: baseline (preintervention), immediately postintervention (Week 2), and at the end of the follow-up period (Week 6). This multiple-time point evaluation allows for a comprehensive analysis of the interventions’ effects on pain intensity, lumbar spine function, quality of life, and anxiety/depression symptoms in patients with LDH.

#### Baseline characteristics of the participants

Data on sociodemographic characteristics, including sex, age, educational attainment, marital status, occupation, economic status, and type of health insurance, will be collected. Clinical and LBP-related information will include body mass index, comorbidities, duration of LBP, and medication use (including type, frequency, and duration).

#### Primary outcome

The primary outcome of this study is pain intensity, which will be assessed via the VAS[18]. The VAS is one of the most widely used instruments for pain evaluation in clinical trials of LBP and has demonstrated high reliability and validity[19]. It consists of a 10-cm horizontal line, scored from 0 to 10, where 0 represents “no pain” and 10 represents “the worst pain imaginable.” The participants were instructed to mark the point on the line that best reflected their average pain level over the preceding 24 hours. Higher scores indicate more severe pain.

#### Secondary outcomes

##### Lumbar Spine Function

The ODI was utilized to assess the impact of LBP on participants’ activities of daily living[20]. The questionnaire comprises 10 items that evaluate functional limitations in domains including pain, lifting, sitting, standing, walking, sleeping, and travelling. Each item is scored on a scale from 0 to 5, with higher total scores indicating a greater degree of lumbar dysfunction.

The JOA score was employed to conduct a multidimensional assessment of LBP. This instrument comprises 25 items across five domains: LBP, lumbar function, walking ability, social life, and psychological status. Higher total scores are indicative of better lumbar functional outcomes.

Three-dimensional gait analysis was performed to comprehensively assess kinematic alterations during walking[21]. Motion data were captured via a six-camera system tracking 22 reflective markers placed on bilateral anatomical landmarks, including the bilateral acromion, greater trochanter, anterior superior iliac spine, ankle, and heel, to record trajectories of the trunk, pelvis, and lower limbs during walking. Representative gait cycles were selected to analyse spatiotemporal parameters—such as step length, walking speed, stride length, and stance phase percentage—along with kinematic parameters, including hip and knee range of motion and hip angular velocity, for the assessment of biomechanical impairments.

sEMG was used to record muscle activity[22]. sEMG electrodes were placed bilaterally over the lumbar erector spinae muscles (ESM), multifidus, and rectus abdominis (RA) of the participants. Specifically, ESM electrodes were positioned at the L1–L3 levels, approximately 3–4 cm from the midline. The multifidus electrodes were located at the L5–S1 level, approximately 2 cm from the spinous processes. RA electrodes were placed 1 cm above the umbilicus and 2 cm lateral to the midline. During trunk flexion-extension tasks, sEMG parameters, including the root mean square (RMS), median frequency (MF), flexion‒relaxation ratio (FRR), and extension‒relaxation ratio (ERR), were calculated. Furthermore, synchronized with three-dimensional gait analysis, dynamic muscle activation patterns were recorded during walking. The gait cycle was segmented into four phases: right double-limb support, left swing, left double-limb support, and right swing. The RMS and MF of the sEMG signals were computed for each phase. The cocontraction index between the ESM and the RA was also assessed to evaluate muscular coordination in LDH patients[23].

##### Pain intensity

EEG was used to identify power in distinct frequency bands (θ, α, β, γ) for monitoring changes in pain sensitivity and pain intensity in participants. Both resting-state and task-state EEG signals were collected. For resting-state EEG, participants were instructed to remain quiet with their eyes open for 3 minutes. For task-state EEG, mildly painful stimuli were applied five times each to the lower back and the dorsum of the right hand. Previous studies have demonstrated an association between EEG patterns and subjective pain perception, supporting the use of EEG as an objective biomarker for assessing pain status in patients with low back pain[24].

##### Quality of life

Quality of life was assessed via the SF-36[25]. This instrument evaluates health-related quality of life across eight domains: physical functioning, role physical, bodily pain, general health perception, social functioning, role emotional, and mental health. Higher scores indicate a better quality of life.

##### Anxiety and depression

The HADS was utilized to assess the risk and severity of anxiety and depression in participants. This instrument effectively evaluates psychological comorbidities frequently associated with pain[26]. The scale consists of 14 items distributed across two subscales: anxiety and depression. For each subscale, a total score of 0–7 is generally classified as normal, 8–10 as borderline or suggestive of possible anxiety/depression, and scores above 10 are indicative of clinically significant emotional disorders.

##### Safety assessment

The incidence of adverse events served as the safety evaluation indicator in this study. Adverse events, such as the emergence of LBP or exercise intolerance, were defined as any intervention-related incidents that occurred during the trial. In the event of an adverse event, the investigator assessed whether the participant could continue the trial. If pain or underlying conditions showed persistent exacerbation, appropriate treatment was promptly initiated, and trial discontinuation was considered. All adverse events were documented and managed promptly by the intervention practitioners, with the overall incidence calculated upon trial completion.

### Data management and confidentiality

Data from scale assessments were collected using case report forms and subsequently entered into an electronic data capture system for real-time entry and monitoring. Objective assessment data were directly acquired and recorded by electronic devices. All data entry personnel received professional training and adhered to a double-data entry verification process to ensure accuracy. To ensure confidentiality, all personally identifiable information was replaced with a unique study identifier, with access restricted solely to the data monitoring committee, data analysts, and research investigators.

### Statistical analysis

Statistical analyses were performed via IBM SPSS Statistics (Version 27.0). A two-sided test with a p-value < 0.05 was considered statistically significant in all analyses. Normally distributed continuous variables are presented as the means and standard deviations, whereas nonnormally distributed variables are summarized as medians and interquartile ranges. The baseline characteristics of the Daoyin group and the CSE group were compared via independent samples t-tests and chi-square tests. The generalized estimating equation (GEE) was employed to assess the effects of group, time, and the group-by-time interaction on outcome variables to determine whether intervention effects varied over time or between groups[27]. The GEE accounts for the longitudinal nature of the data and adjusts for potential confounding factors[28]. Furthermore, depending on the data distribution, either Pearson or Spearman correlation coefficients were used to explore correlations between EEG parameters and VAS scores, as well as between sEMG/three-dimensional gait analysis parameters and ODI/JOA scores.

This study adhered to the intention-to-treat (ITT) principle, whereby all randomized participants were analysed according to their original assignment to minimize bias associated with simplistic handling of protocol deviations[29]. Missing data were addressed via multiple imputation.

## Discussion

This study aimed to compare the effects of Daoyin and CSE on pain intensity, lumbar spine function, quality of life, anxiety, and depression in patients with LDH. The Daoyin intervention integrates static postures with dynamic movements, emphasizing the combination of physical strength and meditative focus[28]. It is performed alongside rhythmic breathing, reflecting the principle of unifying the body, breath, and mind. The consistent and long-term practice of Daoyin may alleviate muscle tension, enhance muscular strength and physical function, improve activities of daily living, and positively affect mental health. However, most previous studies have employed traditional mind–body exercises such as Baduanjin or Tai Chi, which are general-purpose in nature and not specifically tailored to the clinical characteristics of LDH patients[30–32]. Consequently, the specific efficacy of Daoyin in LDH remains uncertain[28,32]. In this study, the selected Daoyin exercises were customized for patients in the chronic stage of LDH. These exercises are designed to improve lumbar flexibility and mobility while strengthening the lumbar and lower limb muscles, thereby alleviating LBP and reducing the risk of recurrence.

Long-term adherence to Daoyin training may enhance lumbar stability and flexibility in patients with LDH, thereby reducing the risk of recurrence. Thus, improving exercise adherence and motivating patients to actively engage in Daoyin practice are essential. The TGA serves as an interactive theoretical framework designed to facilitate behavioral change. Through structured communication between nurses and patients, TGA helps identify barriers, set collaborative goals, and promote ongoing interaction and feedback. Within this process, nurses first establish effective communication to engage patients in shared decision-making. Patients are encouraged to express their concerns and difficulties, which helps them recognize gaps in self-management and fosters motivation for behavioral change. Nurses and patients then work together to establish goals and formulate actionable plans for health promotion. By increasing awareness of unhealthy habits or misconceptions, the TGA enhances patients’ understanding of Daoyin and improves their adherence to home practices. On the basis of this rationale, we integrated the TGA framework with Daoyin exercises for LBP, developing a TGA-based Daoyin intervention protocol. This approach effectively encourages patient participation in Daoyin practice, ensures proper implementation of the intervention, and maximizes the therapeutic benefits of Daoyin for LBP.

Pain is a multidimensional subjective experience influenced by a range of factors, such as tissue injury, sensory perception, and emotional state, which complicates its assessment[24]. In clinical research, pain is commonly measured via patient-reported instruments such as the Numerical Rating Scale or VAS. These tools allow patients to rate their pain on various dimensions, including intensity, duration, and emotional impact[33]. Consequently, the use of such scales has inherent limitations in comprehensively capturing the complex nature of pain.

EEG, a noninvasive neurophysiological monitoring technique, reflects real-time cerebral dynamics by measuring postsynaptic potentials[24]. Numerous studies have employed EEG to evaluate pain, demonstrating correlations between pain perception and oscillations in the θ, α, β, and γ frequency bands[34,35]. Research by Paul et al. indicated that patients with chronic pain exhibit greater θ and β oscillatory power than healthy controls do[36]. In individuals with LBP, pain severity is negatively correlated with α-band power, whereas persistent pain levels are positively correlated with β and γ oscillations in the frontal region[24]. Therefore, we utilized EEG as an objective biomarker to comprehensively evaluate changes in pain by analysing subjects’ EEG characteristics before and after intervention, along with correlation analysis between EEG and VAS scores, thereby determining the effectiveness of the intervention.

Parameters derived from sEMG—such as amplitude, FRR, and cocontraction—together with gait analysis can serve as objective measures reflecting clinical changes in LBP patients. Previous studies have confirmed that, compared with healthy controls, patients with LDH exhibit significantly reduced sEMG amplitudes in the multifidus muscle during trunk extension[22,37]. A diminished lumbar FRR during flexion has also been empirically linked to LBP[37]. Furthermore, research has demonstrated that LDH patients display increased antagonistic trunk muscle cocontraction during both flexion and extension tasks[23]. Altered gait represents another key characteristic of LBP. Studies have shown that individuals with LBP exhibit significant differences in spatiotemporal parameters, impaired pelvic coordination, and abnormal activation of the ESM during walking, often manifesting as reduced walking speed and shortened stride length[21,38]. Based on this evidence, the present study employed three-dimensional gait analysis combined with sEMG to quantitatively assess spatiotemporal gait parameters and the contraction and coordination of corresponding lumbar and abdominal muscles during walking and forward flexion tasks.

This protocol describes the design of an RCT aimed at evaluating the efficacy of Daoyin exercise for LBP in patients with LDH with respect to pain intensity, lumbar function, quality of life, and anxiety/depression symptoms. The study incorporates a rigorous methodological design to ensure the collection of high-quality data. The findings from this trial may offer a solid evidence base to inform clinical practice. Furthermore, the results are expected to help establish a foundation for the clinical management of LDH and provide guidance for future research on Daoyin as a therapeutic intervention for LDH or other conditions.

## Data Availability

No datasets were generated or analysed during the current study. All relevant data from this study will be made available upon study completion.

## Authors’ contributions

Conceptualization: Mutong Wang, Yang Zhang

Funding Acquisition: Yang Zhang

Methodology: Shuai Guo, Yican Yang, Gengyu Liang, Zishan Wang

Writing-Original Draft Preparation: Mutong Wang

Writing-Review & Editing: Mutong Wang, Shuai Guo, Yican Yang, Gengyu Liang, Zishan Wang, Yang Zhang

## Acknowledgements

The authors would like to acknowledge all the members of the Rehabilitation and Physical Therapy Department, Shandong University of Traditional Chinese Medicine Affiliated Hospital.

## Supporting information

S1 Table. SPIRIT 2025 Checklist

S2 Table1. An Intervention Protocol for Low Back Pain (LBP) Daoyin Exercise Based on the Theory of Goal Attainment

S2 Table2. Core Stability Exercise (CSE) Action Specific Steps

